# Cluster randomized controlled trial to assess the effectiveness of a package of community-based intervention on continuum of maternal and newborn healthcare in Sidama, Ethiopia:The SiMaNeH trial protocol

**DOI:** 10.1101/2024.09.01.24312899

**Authors:** Achamyelesh Gebretsadik, Yemisrach Shiferaw, Hirut Gemeda, Yaliso Yaya

## Abstract

**Background:** Maternal and newborn mortality and morbidity remain high in low-and middle-income countries such as Ethiopia. Limited access and dropouts from essential continuum of care interventions are critical factors. In Ethiopia about one in five complete the continuum of essential care through pregnancy, childbirth, and postnatal period Ethiopia. Evidence is limited on whether a package of interventions involving key community health actors increase the proportion completing essential maternal and newborn healthcare continuum in rural Sidama regional state, Ethiopia.

**Objective:** This study aims to implement and evaluate the effectiveness of community-based interventions designed to enhance involvement of key community health actors to improve completion rate of continuum of maternal care and utilization of newborn care.

**Methods:** Twenty rural kebeles (clusters) in Sidama Regional State, Ethiopia, are randomly allocated to intervention and control arms. A total of 2000 pregnant women, 1000 per arm, will be recruited between 20^th^ and 26^th^ week of gestation after intervention. Then they will be followed until six weeks postpartum between June 2024 and February 2025. In the intervention arm, mothers and newborns will receive targeted interventions at home and in their community through a package of interventions designed to improve completion rate of recommended maternal and newborn care. Control clusters will receive normal care from the state public health system. Primary outcomes will be the difference in the completion of continuum of maternal care and essential and emergency newborn care between intervention and control clusters measured by composite indicator constructed from variables. Secondary outcomes include rates of antenatal care completion, facility deliveries with skilled care, completion of at least four postnatal care, essential newborn care, and emergency identification and referrals, mortality measures.

**Conclusion:** This trial will implement and evaluate community-based intervention package within existing community healthcare infrastructure to produce evidence for informed policy and practice to achieve improved community-based healthcare.

## Introduction

Global maternal and child mortality was remarkably reduced between 2000 and 2015 through efforts under Millennium Development Goals (MDGs). However, recent estimates by UN inter-agency highlight that the progress stagnated between 2016 and 2020 (1). In 2020, there were over 300,000 maternal deaths, 2.4 neonatal deaths, and another 1.9 stillbirths (2, 3). Furthermore, neonatal mortality, the death of a newborn baby within the first month of life, shares nearly half (47%) of mortality of under-5 children shows slow progress.

Over 99 percent of maternal and newborn deaths and severe morbidities and disabilities from pregnancy and childbirth occur in low-income and middle-income countries. Unfortunately, sub-Saharan Africa has the highest neonatal mortality rate in the world with 27 per 1000 live births (4), also contributing close to half (42%) of global stillbirths, the death of babies after 28^th^ weeks of gestation and before birth, in 2019 (5). Poor investments (6) and limited access to quality maternal and newborn care in resource-limited settings are important factors because one-third of the neonatal deaths occur within the first day and three-forth in the first week after birth (3). Estimates from modeling 75 high-burdened countries highlighted improved coverage of essential maternal and newborn care interventions could avert over two-third of neonatal deaths, one-third of stillbirths, and over half of maternal deaths every year (7) because 85% of newborn deaths are because of fetal asphyxia, preterm birth, intra-partum complications and neonatal infections (7).

Since the WHO’s Alma Ata declaration in 1978 (8), community-based interventions, intended to bring essential healthcare services closer to these hard-to-reach areas have emerged as important alternatives to improve maternal and newborn healthcare. However, the strategies and its success are not uniform across communities and countries.

Ethiopia, like other low-income countries, struggles with limited access to the life-saving interventions for maternal and newborn health to the larger part of its population living in the rural areas, underscoring the need for innovative and sustainable strategies to close the gap in adverse pregnancy and childbirth outcomes.

One such innovative approach is the Ethiopian Health Extension Program (HEP), a flagship national program rolled out in 2003 by Ministry of Health of Ethiopia (9). In each village of about 5,000 population, two women known as Health Extension Workers (HEWs) trained for one year in general health are placed in a satellite health post to provide health promotion and disease prevention services to residents. These women are permanent employee of Ethiopian government with monthly salary.

The Health Extension Program (HEP) has been instrumental in expanding access to primary healthcare services, demonstrating the feasibility and impact of deploying health extension workers (HEWs) in rural settings. From 2012, another voluntary network of about 30 women per village known as women development team (WDT) were introduced to support the works of HEWs, creating a strong network closer to households. Consequently, a systematic review by Yibeltal Assefa and colleagues showed that the HEP has significantly improved essential preventive and health promotion indicators HEP (10). Yet, concerning the skills of the HEWs to provide essential maternal and newborn care, a study identified up to 88% of HEWs had poor knowledge of neonatal danger signs and expressed that they are not confident enough to attend deliveries (11). This underscores the importance of an approach that improves their confidence, skills, and integration with health facilities.

Furthermore, a 2022 article by Tiruneh GT and colleagues demonstrated that overall, only one-fifth (20%) of women who initiated the first antenatal care completed the continuum of recommended essential care that combines completion of antenatal care, skilled birth attendance, and critical postpartum care (12).

One of the challenges in maternal and newborn care continuum in poorly organized health systems is low proportion of women having postnatal contact with competent health professionals. In 2022, WHO published guidelines for postnatal care and recommends at least four such contacts in the key time periods during the first 24 hours, 48-72 hours, one-two weeks and six weeks after birth (13). Without successful completion of postnatal care during these key periods, it is difficult to succeed in improving the continuum of care.

The low rate of completion of essential continuum of care highlights the importance of a community-based maternal and newborn care system that supports the mother, the baby and family through key community health stakeholders such as skilled and motivated community health workers, trained and convinced community opinion leaders and women support groups. A systematic review synthesized evidence from community-based interventions to improve maternal and newborn health by Zohra S Lassi and colleagues in 2015 showed that interventions with strong community involvement showed significant positive results compared to other community-based intervention without community involvement (14). Previous community-based intervention trials in low-resource settings have demonstrated the important benefits of community-based interventions (15–17). However, many of these trials have been limited by a narrow focus of addressing specific factors, overlooking the complex and interconnected nature of the various community-based actors and stakeholders that influence healthcare utilization in rural communities.

Considering these challenges, this protocol outlines a comprehensive approach to conducting cluster randomized rural trials implementing a package of interventions that includes enhancing involvement of several community actors. The current intervention includes training and supervision and evaluate the effectiveness of these interventions in improving the completion of maternal continuum of care and newborn essential care.

Our hypothesis is that the proposed community-based intervention package intended to engage a broad spectrum of key actors, including community health workers, women’s support groups, influential community leaders, local social welfare scheme systems (known as IDIR in Ethiopia), and the linkages between community and health facility services has a potential to improve maternal and newborn healthcare continuum care outcomes compared to control group receiving the routine public health services. Continuum of maternal and newborn care is a continuity of care during pregnancy, childbirth, and postpartum period with coordinated efforts from home through the health facility (18). To achieve improved rate of completion of the continuum of care, we aim to address the complex barriers such as poor knowledge of the importance of initiation and continuation of care until completion of standard care, transport barriers, and family care related challenges when a mother or baby needs to travel to reach care.

## Overall aim

The aim of the proposed trial is to evaluate the collective effectiveness of a package of community-based interventions on improving continuum of maternal and newborn healthcare.

### Objectives

1. To evaluate whether a set of community-based intervention improve completion of maternal continuum of care : A) The overall completion rate with composite result of combination of at least four antenatal care contacts, delivery in a health facility with skilled provider, and at least four postnatal care contacts and completion rates of each three components of the continuum of maternal care. B) Explore the experience of mothers and care providers on the effect of the intervention for maternal care, including the challenges and opportunities.
2. To determine the effectiveness of the community-based strategies on improving utilization of essential and emergency newborn care: A) WHO recommended four elements of essential newborn (immediate and thorough baby drying, skin-to-skin contact, initiation of early breast feeding within first hour, and delayed cord clamping). B) identification and referral of severely sick babies such as babies with danger signs. C) Cost-effectiveness of the intervention on improving maternal and newborn continuum of health care.

## Methods

### Setting

This trial research project will take place in rural areas of Sidama, a regional state in southern part of Ethiopia. Sidama became an autonomous regional state in 2020, separating from the former Southern Nations, Nationalities, and Peoples Regional State (SNNPRS) which is divided into four new regional states between 2020 and 2023. Sidama is situated approximately 400 kilometers south of Addis Ababa with an estimated population of about 4.6 million people in 2023. The capital of the regional state is Hawassa. Still, more than 80 percent of the population lives in rural areas, where agriculture is the primary economic activity and access to quality healthcare services is limited.

In 2018, an empirical study in one of the districts in Sidama showed that fertility was lower than reported from national estimates with crude birth rate of 22.8 births per 1000 population and total fertility rate of 2.9 children per woman. Furthermore, the population is transitioning to a low-mortality and low-fertility rate (19). However, the United Nations projected crude birth rate for Ethiopia is 30.6 births per 1000 population in 2024 (20), far higher than the empirical findings.

### Theoretical and conceptual frameworks

The trial will employ theoretical and conceptual frameworks, including the WHO-endorsed Participatory Learning and Action (PLA) model (21). The PLA will be used as a guiding model to be used by a team of community resource persons comprised of religious, administrative, and local social scheme leaders, and women support groups. The resource (support) group will use the model to identify, prioritize, address and re-plan based on previous achievements and weakness related to maternal and newborn care in their villages (22). In addition, the intervention will benefit from the process-based approaches of Theory of Change (TOC) principles (23). TOC model helps to understand and implement how activities, outcomes and impacts are connected to each other. Furthermore, WHO and Ethiopian Ministry of health principles of essential maternal and newborn care will be implemented (24, 25) to improve the continuum of care.

### Time Frame

Figure 1 attached in a separated document.

**Figure 1.**
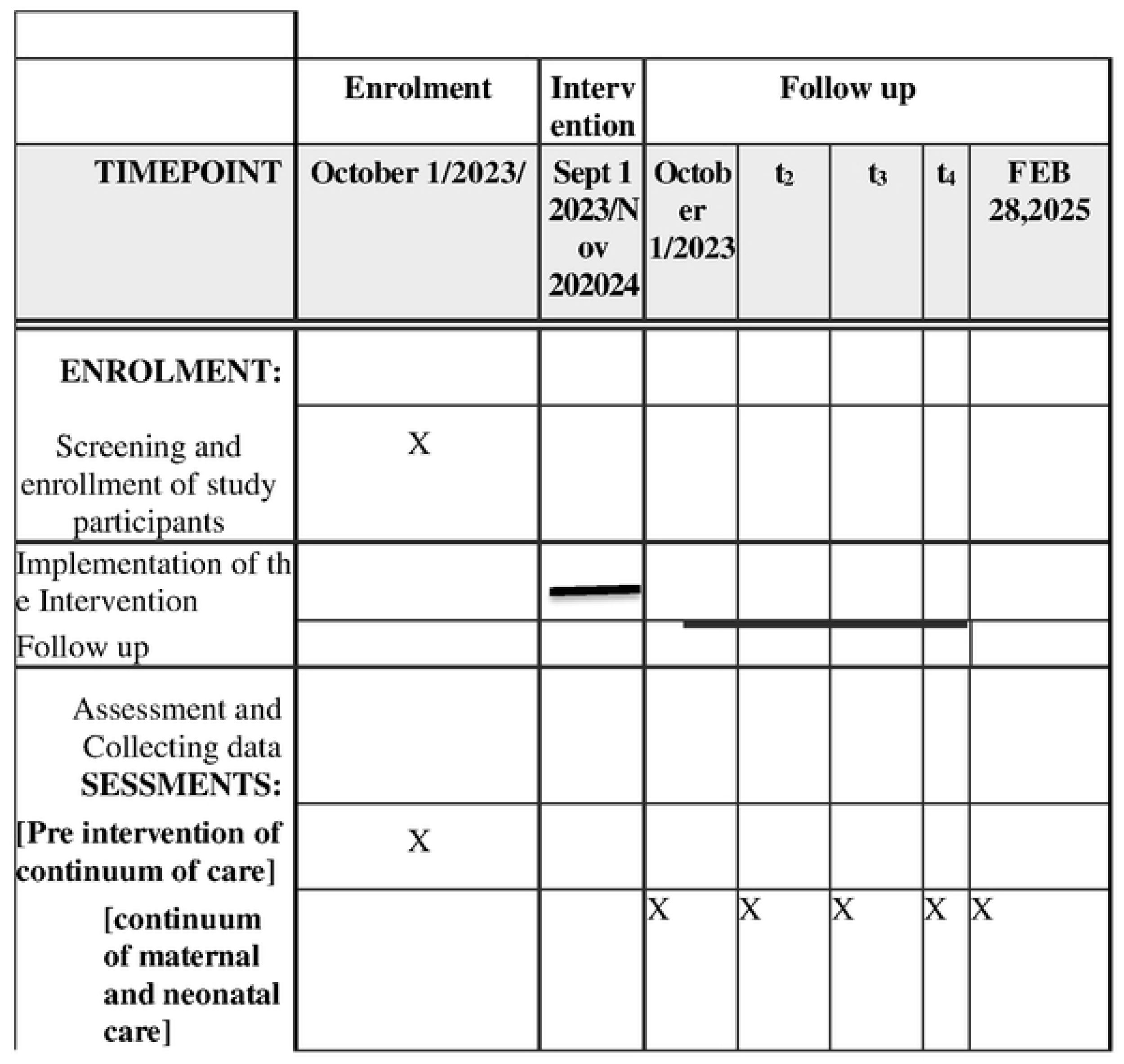
Schedule of enrolment, interventions, and assessments of cluster randomized controlled trial to assess the effectiveness of a package of co1nmunity-based intervention on continuum of maternal and newborn healthcare in Sidama

**Figure 2:**
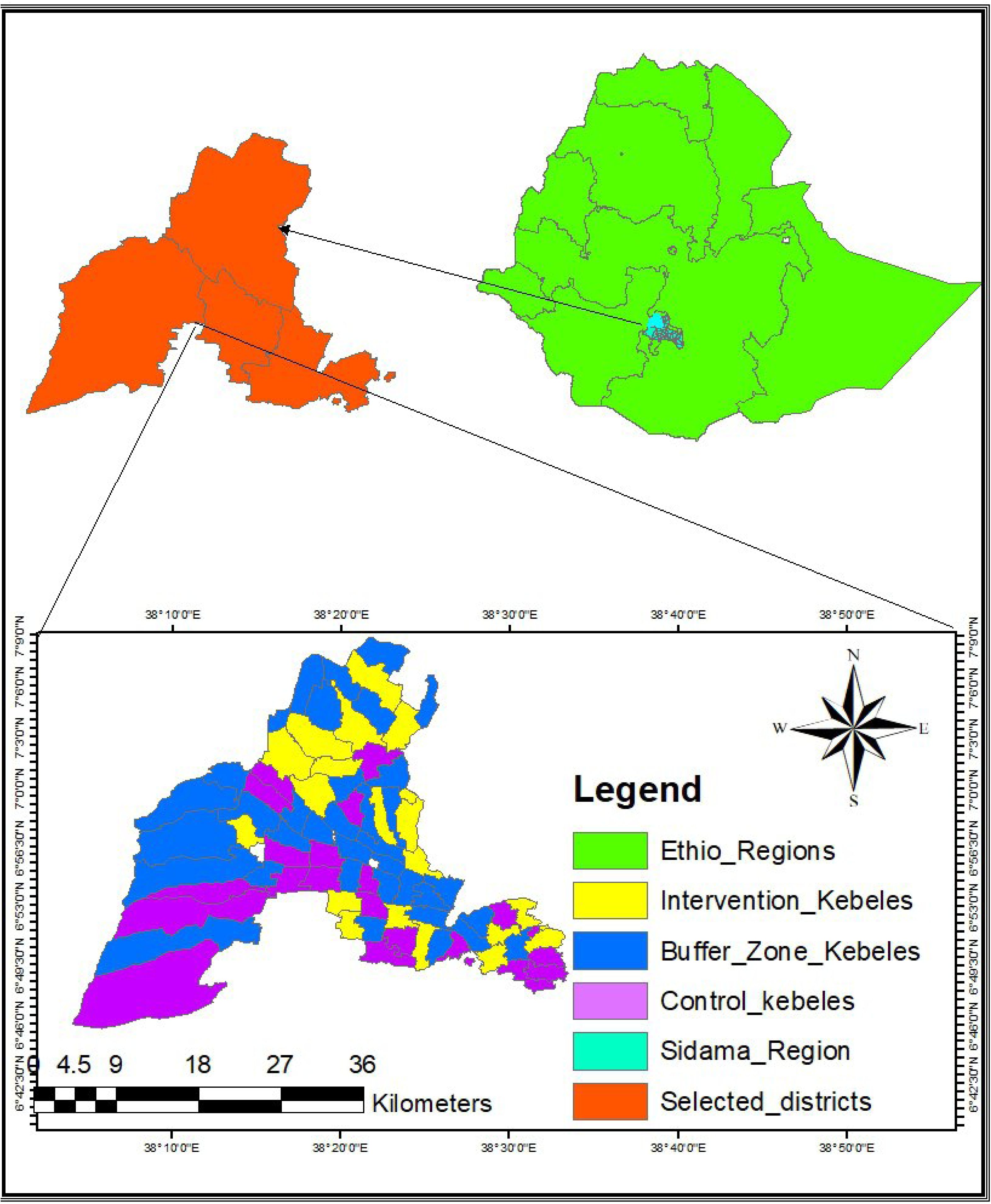
The map of the study area within Sidama regional State, Ethiopia.

**Table 1:** Background demographic and health service characteristics of study population.

| Background | Intervention | Control | Remarks |
| --- | --- | --- | --- |
| Total population in 2024 | 84 347 | 87 830 |  |
| Expected births in 2024<sup>x</sup> | 2 530 | 2 634 |  |
| Number of existing health centers | 7 | 8 | some overlaps |
| Number of Health Extension Workers | 37 | 35 |  |
| Number of Health Developments Team | 545 | 560 |  |
| Average distance to health centers | 5.3 km | 5,1 km |  |

Note: <sup>x</sup> = annual birth rate (3% of population) multiplied by population

### Ethical approval

The study was approved by the institutional review board (IRB) of Hawassa University in Ethiopia in August, 07, 2023 (IRB/363/15). Amendement and extension of IRB approval given on August /05/2024 to the period of August /06/2025. Permission for the study was received from Sidama Regional State Health Bureau and subsequent local authorities. All participants will be asked to give a written consent information sheet to be presented to them and attached with this protocol. Participants will be informed of the aim of the study and their freedom to refuse or participate and withdraw from the trial freely.

### Study design

This study will use a cluster randomized controlled trial with two arms of equal size. We use villages (kebeles in Ethiopian structure) with an average population of about 5,000 people as the clusters served as units of randomization.

### Randomization technique and Blinding

We selected 20 clusters randomly from eligible 80 from four districts in the region and allocated them into intervention and control in 1:1 allocation,10 each to the intervention and control cluster. To balance based on the distance of clusters from nearby health facilities between intervention and control clusters, we used stratified first the 20 clusters into four categories based on the population size and distance to the nearest health facility to reduce confounding. The four strata were: 1) Higher population (over 4,000 residents) and longer distance (more than an hour of walking distance), 2) Higher population and shorter distance (less than an hour walking distance 3) Lower population (less 4,000 residents) and shorter distance and 4) Lower population and longer distance. Then clusters from each stratum were randomly allocated to intervention and control using a computer-generated random number using simple randomization. A seed number was produced and sent from a university of Bergen as a starting point for random number generation. Blinding is difficult because of the nature of cluster randomized trials, but we created a buffer zone between intervention and control clusters to reduce intervention contamination. Data collectors are independent of the intervention process and do not have formal information on which study arm the cluster to which they collect data belongs.

### Sampling strategy and sample size

To calculate the sample size of the participating mothers and babies in the study, we used the sample size calculation for cluster randomized controlled trials with the fixed number of clusters recommended by Karla Hemming and colleagues (26). We fixed the number of clusters to 20 for logistic and access reasons. We base our estimate of pregnant women with annual crude birth rate of 30 births per 1000 population per United Nations Population Division projection for Ethiopia in 2024 (20). The average pregnant mother-baby pair per cluster was estimated to be 150 during the study period. To estimate the minimum required sample, the trial aims to increase the completion rate of the continuum of pregnancy, birth, and postpartum care from an estimated 20% exiting status to 30% in intervention clusters with a 95% significance level and 80% power. We take the intra-cluster correlation of 0.01. Substituting these numbers in the following formula gives the minimum number of mother-baby pairs for the study. We used the following formula to calculate the sample size.

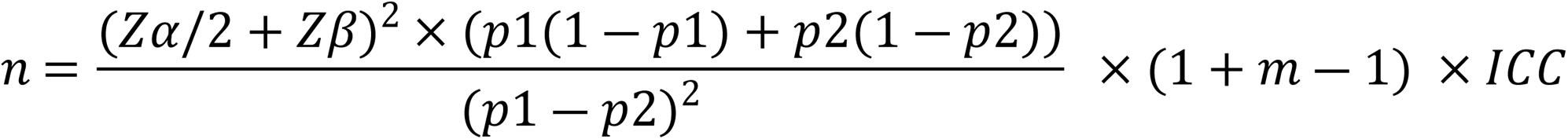

- *n* is the sample size = number of pregnant mother-baby pairs needed per study arm
- *Z_α/2_* is the z-value corresponding to the significance level (α=0.05, Zα/2 =1.96).
- *Z_β_* is the z-value corresponding to the desired power (β = 0.20, Zβ = 0.84).
- *p_1_* is the expected proportion in the control group (0.20), the continuum of care completion in control clusters.
- *p_2_* is the expected proportion in the intervention group (0.30), expected continuum of care completion in intervention clusters.
- *m* is the average cluster size = 150 mothers per cluster on average.
- *ICC* is the intra-cluster correlation coefficient (0.01).

This provides a minimum of 723 pregnant women-baby pairs per trial arm, resulting in a total sample size of 1446. However, because several outcome measures may need more samples and to have adequate numbers for sub-analysis, we decided to recruit a total of 2000 woman-baby pairs, 1000 in each arm.

### Participants

#### Inclusion criteria

We shall include mothers who permanently reside in the study clusters and consent to participate. All pregnant mothers who fulfill the above criteria and do not meet the following exclusion criteria will participate. This includes mothers in their gestational period of between the 20th and 26th weeks within the four months of the recruitment period (June-September 2024).

#### Exclusion criteria

We will not recruit mothers with less 20th weeks of gestation because of longer time needed for follow-up. In addition, mothers whose pregnancy terminated before 20th week of gestation will not be included because the main emphasis of the study is on completion of maternal care and utilization of newborn care. Mothers who do not consent to participate in the study after information will not be included.

### Intervention

To achieve the aim of improving the completion rate of the continuum of essential care from pregnancy care through postnatal care, the intervention package uses four mechanisms of action to help reduce barriers to essential healthcare utilization. The four mechanisms of action are: 1) Actively identify and connect to healthcare services, 2) Educate and prepare at home, 2) Support emotionally and materially through local collective efforts, and 4) Provide home-based essential care in case when care from health facilities is not actual.

To implement the four mechanisms of action stated above, the project coordinates the efforts of three existing community health stakeholder groups through training and supervision. The three stakeholders are the community health workers (HEWs in Ethiopia), the women support groups (HDTs in Ethiopia), and the community resources groups, also termed community opinion leaders (COLs).

HEWs are women permanently employed on average two in all clusters (Kebeles) in Ethiopia and paid monthly salary from the government. They have overall responsibility in their clusters for health promotion, disease prevention, and provision of basic maternal and newborn care such as antenatal, postnatal, and baby resuscitation. HEWs will participate majorly in three of the four mechanisms of action, namely identification, educating and preparing, and provision of home-based essential care when delivery happens at home, in addition to routine postnatal care. Furthermore, they will coordinate the work of COLs for the local collective support efforts.

WDTs are about 30 voluntary groups of women in each cluster residing in sub-villages of clusters in Ethiopia, freely helping the works of HEWs. They have recognition from the government without getting paid. Because of their unique position closer to sub-villages, they have the unique opportunity to identify mothers as they become pregnant in their respective villages and connect to healthcare services. They also educate and prepare mothers and families for standard and emergency care. WDTs’ main contribution to the intervention is actively identifying and connecting mothers and babies to essential standard care and emergency care when they observe danger signs. WDTs will give particular attention to high-risk and hard-to-reach mothers and newborn babies such as extremely poor, living in difficult topography and distant villages from the health facilities and HEWs stations as well as those with existing sicknesses and malnutrition. As such, the contribution of the village WDTs in the four mechanisms of action will be identifying and connecting, education and preparing and involving with COLs in strengthening financial, transport, and family care support through collective local efforts so that a mother or baby reaches to health facility and receives care.

Community opinion leaders (COLs) are key leaders who lead opinions such as approval of the importance of receiving healthcare services and coordinating local collective help mechanisms. COLs are a group of local leaders forming a committee which includes influential religious leaders, cultural leaders, local government administrative leaders, and leaders of local welfare scheme called IDIR in Ethiopia. IDIR is a local welfare system where families contribute financially or materially every month. This welfare system often shares serious costs during deaths and severe sicknesses.

The main responsibility of the COLs is identifying problem such as barriers hindering mothers and babies receiving important health care and fall out of the continuum of care. They respond to the problem coordinating local collective efforts, for example transport or financial support to reach a health facility and taking care of the family members at home. For this contribution, they meet every two-weeks to evaluate and re-plan their actions based on the principles of Participatory Learning and Action (PLA) model (21). WDTs help COLs with gathering and providing sub-village information. COLs use their high level of acceptance to visit and support families where the relationship between the woman and her husband is not smooth and put great pressure on the health and wellbeing of women or their newborns. At such households, COLs provide essential advice on the importance of care for a pregnant women and newborn babies. This is expected to contribute to a mother and baby receive essential care that improve their survival and wellbeing.

The project provides training and supervision support to the three key stakeholders participating in intervention.

For COLs, the training and supervision will be based on PLA model of problem identification, providing local solutions, evaluating the effect, and re-planning process. COLs receive a day-long initial training and a two-monthly supervision discussion for an hour during the project period.

Trainings and supervision to HEWs will have two dimensions: On one hand, HEWs will be trained and supported on technical refresher issues related to baby resuscitation and home-based immediate support when a birth occurs at home, and essential postnatal maternal and newborn care and on the other hand, they will be trained on leadership role to lead the efforts of COLs and WDTs. HEWs receive two days of intensive training followed by two-monthly supervision and monitoring meetings with the project team.

The training and supervision of WDTs will be mainly on identifying and connecting mothers and babies for normal essential care as well as detecting danger signs and referring for emergency care babies and mothers with severe sickness (danger signs). In addition, WDTs receive training on maternal essential self-care, preparation for healthcare, nutrition, rest, hygiene, and other important information which in turn they will teach mothers and families. Further, the WDTs receive training on tracking and giving special attention to high-risk mothers and babies in difficult families and hard-to-reach villages so that they are not left out or fall out of the critically needed healthcare services. Through these mechanisms, the project builds a supportive network around mothers to help improve the initiation and completion of essential services in the continuum of care around pregnancy and childbirth. Table 2 below describes the roles of each stakeholder to the four mechanisms of actions to improve continuum of care in the intervention clusters.

**Table 2:** The role of stakeholders in the community-based actions in intervention clusters.

|  | 1) Identify and connect | 2) Educate and prepare | 3) Support through collective efforts | 4) Home-based essential care |
| --- | --- | --- | --- | --- |
| HEWs | All pregnant mothers for essential ANC, delivery, and postnatal care in collaboration with HDTs and COLs<br>Mothers and babies in high-risk of falling out of care, or even do not initiate first contact.<br>Screen and recruit mothers for study 20-26<sup>th</sup> week of gestation. | Mothers and families on essential care, nutrition, hygiene...<br>Prepare mothers and family for facility delivery, emergency health seeking. | Mainly through coordinating the efforts of COLs | Encourage antenatal care as per the guidelines of MoH<br>Emergency maternal and newborn help for deliveries occurred at home.<br>Routine postnatal care for mothers and babies. |
| HDTs | Identify mothers and babies in her sub-village and connect to HEWs, COLs and health care.<br>Guide data collectors to locate homes. | Educate and prepare mothers and families on essential care, nutrition, hygiene, emergency care | Inform to COLs and HEWs about mothers and babies not receiving care | Educate and prepare.<br>Do not provide any technical care. |
| COLs | Identify mothers and babies at high-risk and hard-to-reach villages.<br>Identify mothers and babies in families with non-smooth relationships and in dangers.<br>Connect those identified to healthcare service. | Approve and promote the importance of essential health care.<br>Make PLA discussion to prepare for support when needed. | Facilitate finance, transport, and family care coordinating local collective efforts.<br>Continuously monitor their support provision | Make home visits to difficult families and provide advice and emotional support. |

*Note: HEWs = Health extension workers, HDTs = Health development team, COLs = Community opinion leaders (religious, cultural, local welfare scheme, and administrative leaders working as a group to ensure support mechanisms), PLA = Participatory Learning and Action (a community problem solving model)*

### Outcome and process indicators

The primary maternal outcome measure is the completion rate of essential maternal care. We define a continuum of care completion as follows:

1. fully completed care when a mother has received four or more antenatal care, delivered in a health facility with a skilled attendant, and received at least four postnatal care (first 24 hours, 48-72 hours, 7-14 days, and six weeks after delivery) from HEWs or other health professionals within the first month after delivery. Because maternal completion of these essential care components will have an implication for newborn health and survival, and essential newborn care is given simultaneously during delivery and postnatal care, the completion of essential newborn care will be analyzed in relation to the maternal completion rate.
2. Emergency home-based care when the delivery occurred at home and mother and baby received immediate visit at home by HEWs or a higher health professional.
3. Sub-analysis on the primary outcome will compare intervention with control clusters separately on antenatal care completion rate, rate of facility delivery, proportion of assistance received immediately form HEWs or other professionals when a birth occurred at home, and proportion of mothers who stayed at maternal waiting shelters immediate days before delivery.
4. Proportion received financial, transport, and family care support from community resource persons (COLs) when a support is critically needed.
5. The proportion of newborn babies received the four essential newborn care services (immediate and thorough drying, skin-to-skin contact, immediate breastfeeding initiation, and delayed cord clamping)
6. Emergency newborn sickness identification and referral rates.
7. Secondary outcome measure will be on whether the intervention affected reduction of neonatal mortality and stillbirth rates.

For newborn babies, we will measure and compare with the control arm the proportion of babies immediately put to breastfeeding and exclusively feeding until at least a month after birth, the proportion of babies received essential thermal care (wrapping in close, skin-to-skin contact to mother immediately, delayed baby bathing), baby resuscitation at home and in clinics, and proportion of babies received emergency treatment and referrals when they had danger signs.

### Data collection

Data will be collected by trained independent data collectors a maximum of five times at households and from health facility records.

First, we will collect data when the mother’s gestation is 20th to 26th week when she formally recruited for subsequent follow-up. This period matches with the time when a mother became pregnant after the start of the intervention process in the intervention cluster. In other words, the mothers who enter the first data collection and follow-up phase on their 20th to 26th week of gestation has been exposed to interventions since they were pregnant. In this first round of data collection, data will be collected on basic socio-demographic and important variables for important covariates such as wealth status indicators, distance to health facility, history of healthcare utilization during the previous pregnancy and delivery.

Follow-up data collection will include the second to the fifth rounds of data collection. The second round will be 10 weeks after the first data collection (30^th^ to 36^th^ week of gestation). This data collection will mainly focus on information about variables related to the continuation of antenatal care utilization and whether the mother had pregnancy related complications and severe sickness, including information on received healthcare for sickness.

The third round of data collection is on the fourth day after delivery. The main variables in this round of data collection include variables on antenatal care completion, delivery information such as place of delivery, delivery attendant, utilization of postnatal care during the WHO recommended first 24 hours and 48-72 hours after birth, whether the mother received any financial, transport, or family care support from the COLs support system to reach to health facility, whether the mother stayed in the maternal waiting shelter nearby health facilities the immediate days before delivery, delivery outcome (livebirth or stillbirth). Additional information will include about maternal and newborn complications during delivery, essential newborn care provided during delivery (resuscitation, umbilical care, thermal care, initiation of breastfeeding, treatment for severely sick and premature babies).

The fourth round of data will be collected two weeks after delivery (10 days after the third data collection) on key first week survival, first week occurrence of morbidity, emergency help received for those who had serious sickness and utilization of postnatal care during the WHO recommended 7-14 days after birth, and essential baby care within the first two week.

The final and fifth round of data collection will take place six weeks after delivery. This data collection will include variables on the first month and six week essential postnatal maternal and neonatal care, first month and six-week survival of the newborn, and information on sickness and emergency healthcare received for sickness in this period.

### Data storage and security plan

Data will be transferred from koboCollect, a tablet-based application, directly to the data server at Hawassa University research center. Only people involved in the project will have a password-based access. Data will be de-identified of personal identifiers when stored. Personal variables will never be published anywhere. However, deidentified data will be available to researchers when requested with clear plan for use as stated in the protocol registration.

### Data entry and analysis plan

Data will be double entered into excel sheet and transferred to R software version 4.4.1for analysis (27). Data will be analyzed and presented in a format comparing outcomes between intervention and control arms using intention-to-treat analysis, meaning all recruited will be analyzed irrespective of finishing the study process. Data analysis will use techniques that consider the clustered nature of the data.

### Trial registration

This trial is registered at Pan African Clinical Trial Registry and its number is PACTR202402782261294 and the link is https://pactr.samrc.ac.za/TrialDisplay.aspx?TrialID=25464

### Dissemination plan

We will communicate the result with local government structures, Sidama Regional State Health Bureau and publish scientific articles in internationally recognized peer-reviewed journals.

### Funding source

Southern Ethiopia Network of Universities for Public Health (SENUPH-II) funds the study through the financial support from Norwegian Programme for Capacity Development in Higher Education and Research for Development (NORHED-II)-Project number 59360. The funders have no role in the study design, data collection, data management and analysis and interpretation of data, writing of the report, and decision to submit for publication.

### Potential strengths and limitations

The primary strength of this trial lies in its innovative approach that implement comprehensive strategies involving a wide range of key community stakeholders. Ethiopian government system has a strong local structure with important stakeholder to support health interventions. These stakeholders include religious, cultural, administrative, and local welfare scheme leaders, who lead and influence opinions and collective resources in rural communities. They will work collaboratively alongside the community health workers and women’s support groups. To improve initiation and continuum in the care, this trial proposes an integrative approach aimed at actively identifying and connecting women and newborns to routine and emergency healthcare services, with increased emphasis on women and newborns in vulnerable households and hard-to-reach villages.

The strategy focuses on strengthening existing local mechanisms to ensure initiation and continuity of essential care. This includes facilitating access to finance, transport, and family care, in addition to emotional support thereby addressing critical barriers to healthcare access.

Despite the strengths described above, the trial may face limitations due to its focus primarily on the quantitative improvement of healthcare utilization, without addressing challenges related to the quality of healthcare. The quality of healthcare measured through quality of provision and the personal experiences of service recipients are crucial factors influencing the continued utilization of healthcare services. However, given the limited scope and resources, the current trial prioritizes the initial step of connecting individuals to healthcare which in turn has the potential to create pressure demanding quality of care. We believe that creating demand through such as out trials may lead to interventions specifically targeting the improvement of healthcare quality.

## Data Availability

Deidentified research data will be made publicly available when the study is completed and published.

## References

1. WHO. Trends in maternal mortality 2000 to 2020: estimates by WHO, UNICEF, UNFPA, World Bank Group and UNDESA/Population Division. Report February, 2023. Accessed April 3, 2024 https://iris.who.int/bitstream/handle/10665/366225/9789240068759-eng.pdf?sequence=1. 2023.

2. Hug L, You D, Blencowe H, Mishra A, Wang Z, Fix MJ, et al. Global, regional, and national estimates and trends in stillbirths from 2000 to 2019: a systematic assessment. The Lancet. 2021;398(10302):772–85.

3. Sharrow D, Hug L, You D, Alkema L, Black R, Cousens S, et al. Global, regional, and national trends in under-5 mortality between 1990 and 2019 with scenario-based projections until 2030: a systematic analysis by the UN Inter-agency Group for Child Mortality Estimation. The Lancet Global Health. 2022;10(2):e195–e206.

4. WHO. Levels and trends in child mortality: report 2021: estimates developed by the UN Inter-agency Group for Child Mortality Estimation. December 12, 2021. Accessed April 3, 2024. https://www.who.int/publications/m/item/levels-and-trends-in-child-mortality-report-2021. 2021.

5. Hug L, Mishra A, Lee S, You D, Moran A, Strong KL, et al. A neglected tragedy the global burden of stillbirths: report of the UN inter-agency group for child mortality estimation, 2020. United Nations Children’s Fund; 2020.

6. Middleton PF. Donor aid and research funding for newborn babies and preventing stillbirths. The Lancet Global Health. 2023;11(11):e1678–e9.

7. Bhutta ZA, Das JK, Bahl R, Lawn JE, Salam RA, Paul VK, et al. Can available interventions end preventable deaths in mothers, newborn babies, and stillbirths, and at what cost? Lancet. 2014;384(9940):347-70.

8. WHO. World Health Organization. Regional Office for Europe. (1978). Declaration of Alma-Ata. World Health Organization. Regional Office for Europe. https://iris.who.int/handle/10665/347879. WHO Europe; 1978.

9. FMOH-Ethiopia. Health Sector Strategic Plan (HSDP-III) 2005/6-2009/10 FMOH; 2005 [Accessed April 3, 2024]. Available from: http://www.nationalplanningcycles.org/sites/default/files/planning_cycle_repository/ethiopia/ethiopia-health-sector-development-planhsdp-iii.pdf. Addis Ababa, 2005.

10. Assefa Y, Gelaw YA, Hill PS, Taye BW, Van Damme W. Community health extension program of Ethiopia, 2003–2018: successes and challenges toward universal coverage for primary healthcare services. Globalization and health. 2019;15:1-11.

11. Birhanu Z, Godesso A, Kebede Y, Gerbaba M. Mothers’ experiences and satisfactions with health extension program in Jimma zone, Ethiopia: a cross sectional study. BMC health services research. 2013;13:1–10.

12. Tiruneh GT, Demissie M, Worku A, Berhane Y. Predictors of maternal and newborn health service utilization across the continuum of care in Ethiopia: A multilevel analysis. PloS one. 2022;17(2):e0264612.

13. Organization WH. WHO recommendations on maternal and newborn care for a positive postnatal experience: World Health Organization; 2022.

14. Lassi ZS, Bhutta ZA. Community-based intervention packages for reducing maternal and neonatal morbidity and mortality and improving neonatal outcomes. Cochrane database of systematic reviews. 2015(3).

15. Bhutta ZA, Soofi S, Cousens S, Mohammad S, Memon ZA, Ali I, et al. Improvement of perinatal and newborn care in rural Pakistan through community-based strategies: a cluster-randomised effectiveness trial. The Lancet. 2011;377(9763):403–12.

16. Midhet F, Becker S. Impact of community-based interventions on maternal and neonatal health indicators: Results from a community randomized trial in rural Balochistan, Pakistan. Reproductive health. 2010;7:1–10.

17. Mushi D, Mpembeni R, Jahn A. Effectiveness of community based safe motherhood promoters in improving the utilization of obstetric care. The case of Mtwara Rural District in Tanzania. BMC pregnancy and childbirth. 2010;10:1–9.

18. Kerber KJ, de Graft-Johnson JE, Bhutta ZA, Okong P, Starrs A, Lawn JE. Continuum of care for maternal, newborn, and child health: from slogan to service delivery. The Lancet. 2007;370(9595):1358–69.

19. Areru HA, Dangisso MH, Lindtjørn B. Births and deaths in Sidama in southern Ethiopia: findings from the 2018 Dale-Wonsho Health and Demographic Surveillance System (HDSS). Global Health Action. 2020;13(1):1833511.

20. UNdata. Statistics: Crude birth rate (births per 1,000 population), Ethiopia, 2024. Available at: https://data.un.org/Data.aspx?d=PopDiv&f=variableID%3A53. Accessed 08 July 2024. UNdata. 2024.

21. Cazottes I, Costello A, Davis J, George A, Houeto D, Howard-Grabman L, et al. WHO recommendation on community mobilization through facilitated participatory learning and action cycles with women s groups for maternal and newborn health. World Health Organization; 2014.

22. Pulkki-Brännström A-M, Haghparast-Bidgoli H, Batura N, Colbourn T, Azad K, Banda F, et al. Participatory learning and action cycles with women’s groups to prevent neonatal death in low-resource settings: A multi-country comparison of cost-effectiveness and affordability. Health Policy and Planning. 2020;35(10):1280–9.

23. Breuer E, Lee L, De Silva M, Lund C. Using theory of change to design and evaluate public health interventions: a systematic review. Implementation Science. 2015;11:1–17.

24. Diego EK, Ehret DE, KC A, Bose CL. Quality Indicators to Evaluate Essential Newborn Care in Low-and Middle-Income Countries. Pediatrics. 2023;152(3).

25. Wojcieszek AM, Bonet M, Portela A, Althabe F, Bahl R, Chowdhary N, et al. WHO recommendations on maternal and newborn care for a positive postnatal experience: strengthening the maternal and newborn care continuum. BMJ Global Health. 2023;8(Suppl 2):e010992.

26. Hemming K, Girling AJ, Sitch AJ, Marsh J, Lilford RJ. Sample size calculations for cluster randomised controlled trials with a fixed number of clusters. BMC medical research methodology. 2011;11:1–11.

27. R Core Team (2021). R: A language and environment for statistical computing. R Foundation for Statistical Computing, Vienna, Austria. Available at: https://www.R-project.org/.

